# Early versus delayed feeding after push-technique percutaneous endoscopic gastrostomy: A randomized controlled trial

**DOI:** 10.64898/2025.12.07.25341795

**Authors:** Thammawat Parakonthun, Wison Ruangsetakit, Chawisa Nampoolsuksan, Nicha Srisuworanan, Atthaphorn Trakarnsanga, Chainarong Phalanusitthepha, Jirawat Swangsri, Asada Methasate

**Affiliations:** Minimally Invasive Surgery Unit, Department of Surgery, Faculty of Medicine Siriraj Hospital, Mahidol University, Bangkok, Thailand; Siriraj Upper Gastrointestinal Cancer Center, Department of Surgery, Faculty of Medicine Siriraj Hospital, Mahidol University, Bangkok, Thailand

**Author notes:** **Corresponding author:** Asada Methasate, MD, PhD, Department of Surgery, Faculty of Medicine Siriraj Hospital, Mahidol University (AM).

**Keywords:** Early, Endoscopic, Gastrostomy, Residual volume, Tube feeding

## Abstract

**Background and Aims:** Traditional guidelines recommend delayed feeding after percutaneous endoscopic gastrostomy (PEG) because of concerns about postprocedural complications. Early feeding is safe after pull-technique PEG; however, optimal timing after push-technique PEG remains uncertain. We evaluated the safety and efficacy of early vs delayed feeding after push-technique PEG, focusing on maximal gastric residual volume, complications, and wound site bacterial colonization.

**Methods:** A randomized noninferiority trial was conducted at a university hospital in 160 patients undergoing successful push-technique PEG placement between February 2022 and June 2023. Patients were randomized 1:1 to early feeding (<4 hours) or delayed feeding (>12 hours). Both groups received identical intermittent drip-feeding protocols. The primary outcome was maximal gastric residual volume. Secondary outcomes were length of stay, opioid consumption, postprocedural complications, and wound culture.

**Results:** Median time to feeding was 2.2 hours (0.7–4.0) in the early group and 17.2 hours (12.8–40.5) in the delayed group. Maximal gastric residual volume did not differ between groups (0 vs 0 mL; *P*=0.26). Early feeding reduced hospital stay (42.3 vs 46.0 hours; *P*<0.01) and opioid use (2.0 [0–10] vs 2.0 [0–12] mg; *P*=0.01). Rates of complications (fever, bleeding, wound infection, abdominal distension) were comparable (20.0% vs 27.5%; *P*=0.35). Wound cultures showed no significant difference in bacterial colonization (26.3% vs 22.5%; *P*=0.58).

**Conclusions:** Early enteral feeding within 4 hours after push-technique PEG is safe, does not impair gastric emptying, and reduces hospital stay and opioid use. These findings extend the evidence for early feeding to push-technique procedures.

**Trial Registration:** Thai Clinical Trials Registry. Registration number TCTR20220105003. Web link: https://www.thaiclinicaltrials.org/show/TCTR20220105003.

## Introduction

Percutaneous endoscopic gastrostomy (PEG), introduced in 1980, is the preferred method for long-term enteral feeding because it is minimally invasive and safer than open surgical techniques. It is commonly indicated for persistent dysphagia from upper gastrointestinal malignancies, head and neck cancers, cerebrovascular diseases, and traumatic brain injuries. Conventional practice delays PEG feeding for at least 24 hours owing to concerns about postprocedural complications; however, growing evidence supports earlier feeding. Multiple meta-analyses found that initiating enteral nutrition within 4 hours after PEG placement does not increase gastric residual volume (GRV) or mortality [1–4]. Randomized controlled trials similarly show no increase in complications with early feeding, and high-acuity populations, including intensive care patients, tolerate early nutrition [5–8]. More recent literature suggests early feeding is not only feasible but also advantageous [9–11]. Studies from general wards, trauma units, and burn centers report comparable safety between early and delayed feeding [12–14]. Current guidelines from the American Society for Gastrointestinal Endoscopy [15], the American Society for Parenteral and Enteral Nutrition [16], and the European Society for Clinical Nutrition and Metabolism [17] endorse feeding within 24 hours. Nevertheless, the optimal initiation time remains controversial and often depends on individual endoscopist judgment [18,19]. Delaying initiation may prolong hospital stay and impede nutritional improvement.

Peristomal infection is another concern after PEG placement. Bacterial colonization of the PEG site occurs in up to 30% of cases, often with skin flora such as *Staphylococcus aureus*, *Pseudomonas aeruginosa*, and *Candida* spp [20–21]. Although prophylactic antibiotics are routinely recommended to reduce infection risk, colonization may still occur, particularly in immunocompromised or critically ill patients. Few studies have examined the incidence of PEG wound infection or microbial colonization at the wound site.

At our center, the push-technique for PEG placement with an intermittent-drip feeding method is standard practice. Most early-feeding studies have examined the pull-technique and included both intermittent and continuous-drip regimens; therefore, optimal timing after push-technique PEG remains uncertain. This study aimed to compare maximal GRV between early feeding (within 4 hours) and delayed feeding (after 12 hours). We hypothesized that early feeding via push-technique PEG is not inferior to delayed feeding with respect to maximal GRV. We also evaluated postprocedural complications, length of hospital stay, opioid consumption, and wound culture results for microbial colonization.

## Materials and Methods

### Study Design and Oversight

We conducted a noninferiority randomized controlled trial in patients undergoing push-technique PEG at our university hospital.

### Sample Size

The sample size was determined using a 2-group *t* test formula for equivalence in means. The standard deviation for GRV was based on a previous randomized controlled trial [9]. The initial calculation required 70 participants per group; to accommodate potential dropouts, we added 15%, yielding 80 per group and 160 total for analysis.

### Participants

From 16 February 2022 to 3 August 2023, we enrolled patients aged 18–85 years who required PEG for esophageal cancer, head and neck cancer, cerebrovascular disease, or traumatic brain injury and were scheduled for push-technique placement. Eligible patients received study information and provided written informed consent. Exclusion criteria were predicted life expectancy < 30 days, body mass index < 18 or > 30 kg/m², and American Society of Anesthesiologists class ≥ 4. We also excluded patients with Eastern Cooperative Oncology Group performance status 4, serum albumin < 3 mg/dL, or anatomical abnormalities visible endoscopically that precluded PEG placement.

### PEG Technique

All PEG insertions were performed under endoscopic guidance using the push technique. Intravenous antibiotics were routinely administered before the procedure for prophylaxis. After transillumination confirmed proper site selection, the skin was prepared with povidone-iodine and infiltrated with 1% lidocaine. Transfascial gastropexy was performed using nonabsorbable sutures. The gastrostomy tube was placed under direct visualization. Any immediate bleeding was coagulated before completion of the procedure. The wound was covered with a dry gauze dressing using sterile technique, and participants were transferred to the recovery room. After 1–2 hours of stable postprocedural monitoring, participants returned to the inpatient ward.

### Randomization and Intervention

All 160 participants were randomized 1:1 by computer-generated simple randomization to early feeding (intervention) or delayed feeding (control). Allocation sequences were kept in sealed envelopes and opened only after PEG insertion was completed by 2 research assistants not involved in intervention evaluation. For early feeding, enteral nutrition was initiated within 4 hours of placement; in the delayed group, feeding began after 12 hours. Only the data analyst was blinded to the assigned intervention. All medications were resumed per standard protocol after feeding initiation, except antiplatelet and anticoagulant therapies, which were resumed 24–48 hours after the procedure if no postprocedural bleeding occurred (**Fig 1**)

**Fig 1.**
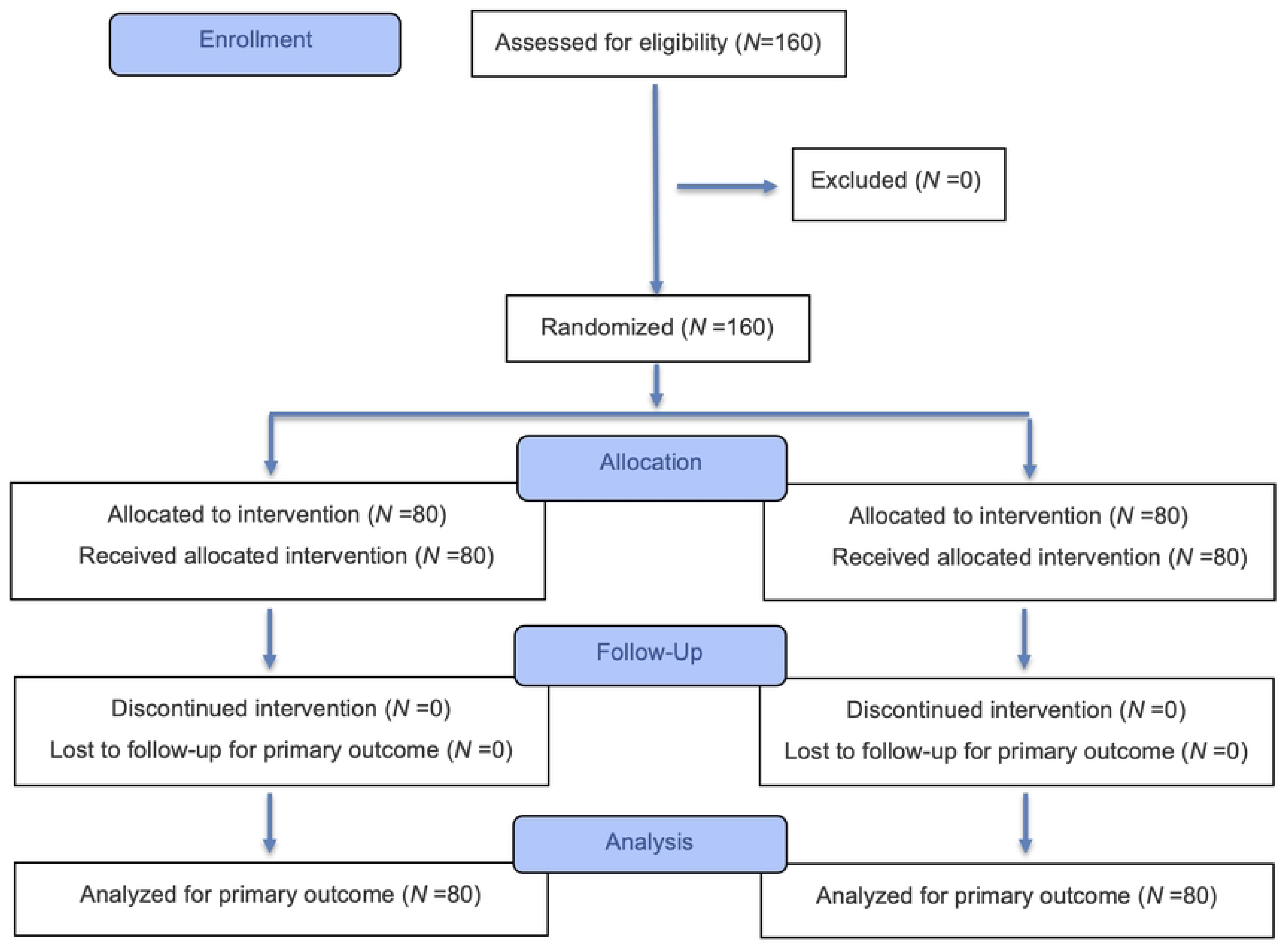
CONSORT flow diagram of participant enrollment, randomization, allocation, follow-up, and analysis. **Abbreviations:** CONSORT, Consolidated Standards of Reporting Trials; *N*, group size

### Feeding Protocol

All participants received the same polymeric formula administered by ward nurses; patients were positioned with head elevated 30–45 degrees during feeding and for 2 hours afterward. A water tolerance test of 100 mL over 30 minutes preceded formula initiation. If tolerated without abnormal symptoms, 200 mL of formula was administered by drip over 2 hours. In the early group, the water test was performed in the recovery room after PEG placement, with the first meal beginning upon ward arrival. In the delayed group, the water test was performed 12 hours after the procedure, followed by formula feeding. Feeding volumes were 200 mL for meals 1 and 2, and they increased to 400 mL for meals 3 and 4. Each meal was followed by 50 mL of water, and meals were spaced 6 hours apart.

### GRV Assessment

The PEG extension kit was connected to the PEG site, and gastric contents were aspirated with a feeding syringe. GRV was measured by ward nurses before each meal. Feeding continued if GRV was < 200 mL or was 200–400 mL without gastrointestinal symptoms; feeding was withheld if GRV was 200–400 mL with symptoms or > 400 mL. The maximal GRV for each participant was recorded after all meals were completed (**Fig 2**).

**Fig 2.**
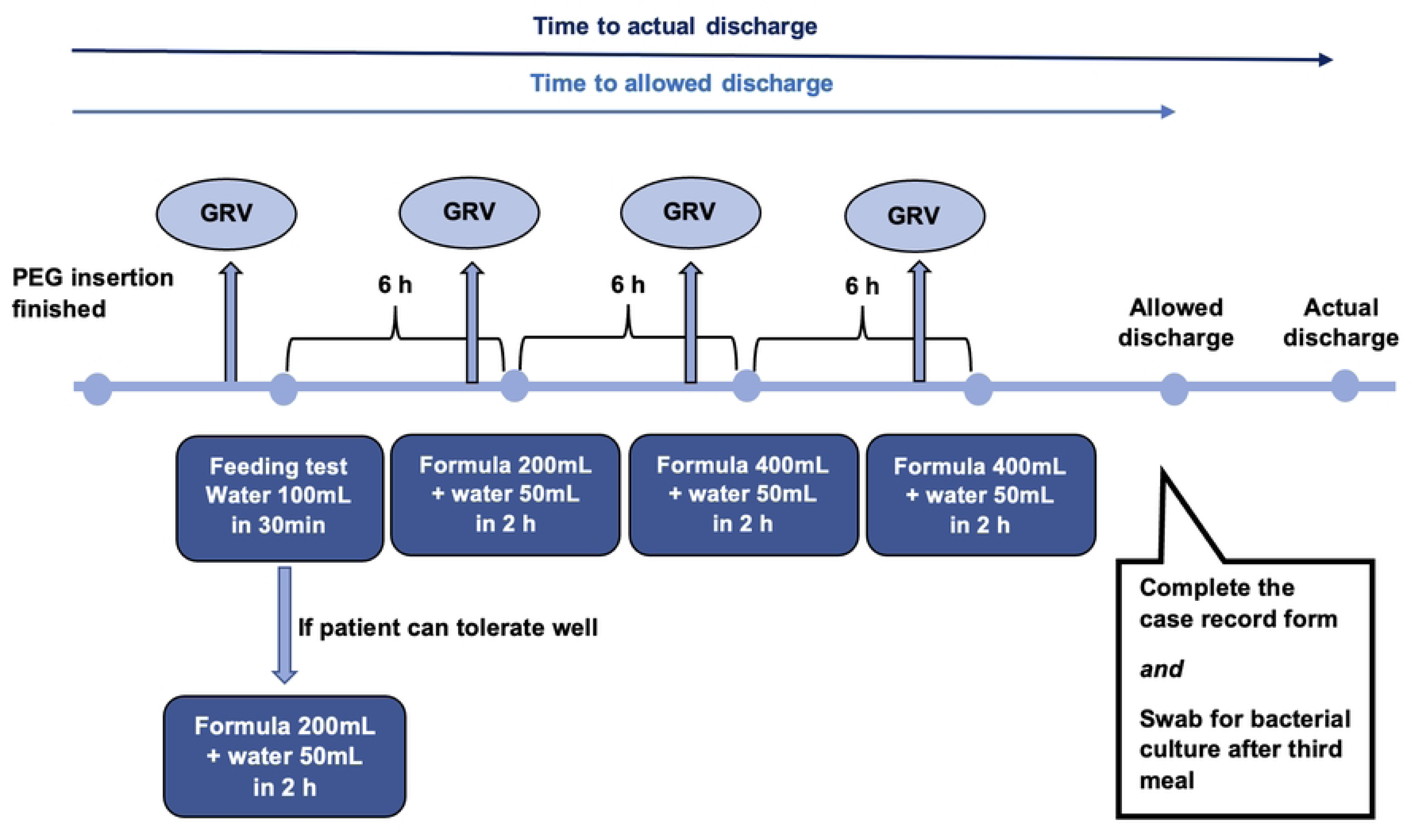
Post-PEG intermittent-drip feeding protocol and outcome assessments. **Abbreviations:** GRV, gastric residual volume; h, hours; mL, milliliters; min, minutes; PEG, percutaneous endoscopic gastrostomy

### Discharge Criteria and Wound Culture

Participants were eligible for discharge when all criteria were met: improved general condition with stable vital signs, and absence of abnormal gastrointestinal symptoms (eg, nausea, vomiting, abdominal distension). Patients had to tolerate all 4 feeding meals, have a caregiver who was capable of home PEG care, and undergo wound swab culture at the PEG wound site after the third meal.

### Data Collection and Statistical Analysis

GRV was recorded for all participants and compared between the 2 groups. Postprocedural variables included initial feeding time, time to allowed discharge, time to actual discharge, total opioid consumption, and complications. Analyses were performed with IBM SPSS Statistics version 26 (IBM Corp, Armonk, NY, USA). Data were reported as *n* (%) or mean (SD) for normally distributed data, or as median (min–max) for nonnormally distributed data. Quantitative variables were compared with the independent *t* test, and qualitative variables with the chi-square test, using the delayed feeding group as the control. A *P* value < .05 was considered statistically significant.

## Results

### Baseline Characteristics

All 160 patients were enrolled, with no exclusions; *N* = 80 were randomized to each group, and there were no intervention discontinuations or missing data. There were no significant between-group differences in sex, age, body mass index, or serum albumin. Men predominated overall, and malignancy was more common than nonmalignant indications; among cancers, head and neck and esophageal cancers were most frequent (**Table 1**).

**Table 1.** Baseline Characteristics and Indications, by Feeding Group.

| Demographic data | Early<br>($N=80$) | Delayed<br>($N=80$) | *P* value |
| --- | --- | --- | --- |
| Male sex | 70 (87.5) | 65 (81.3) | 0.28 |
| Age | 61.9 ± 10.9 | 59.6 ± 11.6 | 0.19 |
| BMI (kg/m<sup>2</sup>) | 19.5 (15.2–29.9) | 19.0 (14.4–29.0) | 0.33 |
| Serum albumin (mg/dL) | 3.7 (2.0–4.8) | 3.8 (3.0–5.0) | 0.39 |
| Disease categories |  |  | 0.55 |
| - Head and neck cancer | 45 | 41 |  |
| - Esophageal cancer | 20 | 24 |  |
| - Other cancers with malnutrition | 3 | 0 |  |
| - Hemorrhagic stroke | 3 | 0 |  |
| - Ischemic stroke | 4 | 3 |  |
| - Traumatic brain injury | 1 | 2 |  |
| - Other neurological disorders | 4 | 9 |  |
| - Benign esophageal stricture | 0 | 1 |  |

Data are presented as *n* (%), mean (SD), or median (min–max). The *P* value for disease categories compares the overall distribution across categories. *N* indicates group size; *n* indicates patients with the characteristic.
**Abbreviations:** BMI, body mass index; SD, standard deviation

### Primary Outcome

Maximal GRV did not differ between groups (0 vs 0 mL; *P* = 0.26). Nonzero GRV occurred in 5 participants in the early group and 3 in the delayed group; none had abdominal discomfort, nausea, or vomiting, and none required cessation of subsequent meals.

### Secondary Outcomes

Median time to initial feeding was significantly shorter with early feeding (2.2 [0.7–4.0] hours) than with delayed feeding (17.2 [12.8–40.5] hours). Early feeding shortened median postprocedural hospital stay (42.3 [20.8–142.1] vs 46.0 [37.5–595.2] hours; *P* < 0.01) and was associated with lower opioid consumption (2.0 [0–10.0] vs 2.0 [0–12.0] mg; *P* = 0.01).

### Complications

Postprocedural complications, including fever, wound bleeding, wound infection, abdominal distension, and nausea or vomiting, occurred at similar rates (20.0% vs 27.5%; *P* = 0.348; **Table 2**). Fever, defined as body temperature > 38 °C, was the most common event; the maximum complication severity was Clavien–Dindo grade 2 in both groups. No deaths occurred.

**Table 2.** Clinical Outcomes After Push-technique Percutaneous Endoscopic Gastrostomy, by Feeding Group.

| **Outcomes** | **Early**<br>**(N=80)** | **Delayed**<br>**(N=80)** | ***P* value** |
| --- | --- | --- | --- |
| **Initial feeding time (h)** | 2.2 (0.7–4.0) | 17.2 (12.8–40.5) | **<0.01** |
| **Maximal GRV (mL)** | 0 (0–200) | 0 (0–260) | 0.26 |
| **Time to allowed discharge (h)** | 22.8 (19.5–26.7) | 37 (22.8–60.5) | **<0.01** |
| **Time to actual discharge (h)** | 42.3 (20.8–142.1) | 46.0 (37.5–595.2) | **<0.01** |
| **Total doses of IV morphine (mg)** | 2.0 (0–10.0) | 2.0 (0–12.0) | **0.01** |
| **Post-procedural complications** | 16 (20) | 22 (27.5) | 0.35 |
| - Fever | 8 | 13 |  |
| - Wound bleeding | 1 | 3 |  |
| - Nausea/vomiting | 4 | 1 |  |
| - Diarrhea | 2 | 2 |  |
| - Pneumonia | 1 | 1 |  |
| - Congestive heart failure | 0 | 1 |  |
| - Abdominal distension | 0 | 1 |  |
| - Wound infection | 0 | 1 |  |
| - Urinary tract infection | 0 | 1 |  |
| **Positive bacterial culture results** | 21 (26.3) | 18 (22.5) | 0.58 |

Data are presented as *n* (%) or median (min–max). “Time to allowed discharge” is the earliest time all discharge criteria were met. “Time to actual discharge” is the recorded hospital discharge time. Fever is defined as body temperature > 38 °C. Comparisons used the independent *t* test for continuous variables and the chi-square test for categorical variables. *N* indicates group size; *n* indicates patients with the characteristic.
**Abbreviations:** GRV, gastric residual volume; h, hours; IV, intravenous

### Wound Culture

Bacterial colonization on wound culture did not differ between groups (26.3% vs 22.5%; *P* = 0.58), with coagulase-negative *Staphylococcus* most frequently isolated (**Table 3**).

**Table 3.** Peristomal Wound Culture Results, by Organism and Feeding Group.

| **Bacterial culture** | **Early**<br>**(N=80)** | **Delayed**<br>**(N=80)** |
| --- | --- | --- |
| Coagulase-negative *Staphylococcus* | 14 | 10 |
| *Staphylococcus aureus* | 2 | 1 |
| *Klebsiella pneumoniae* | 0 | 2 |
| *Micrococcus* spp | 0 | 2 |
| *Pseudomonas aeruginosa* | 0 | 2 |
| *Bacillus* spp | 1 | 1 |
| *Escherichia coli* | 1 | 1 |
| *Citrobacter* spp | 0 | 1 |
| *Stenotrophomonas maltophilia* | 1 | 0 |
| *Corynebacterium* spp | 1 | 0 |
| *Acinetobacter baumannii* | 1 | 0 |
| *Candida* spp | 1 | 0 |

Counts reflect organisms isolated from peristomal wound swab culture performed after the third meal. Totals may exceed the number of positive cultures because some specimens yielded multiple organisms.
**Abbreviations:** PEG, percutaneous endoscopic gastrostomy; spp, species

## Discussion

Since its introduction in 1980, PEG has become a widely used method for long-term enteral feeding in appropriately selected patients. Its minimally invasive nature, technical feasibility, and low complication rate have made it preferred over traditional surgical gastrostomy. PEG is particularly beneficial for patients with neurological disorders, head and neck or esophageal malignancies, and dysphagia or malnutrition. Advances in technique and equipment have further improved safety and broadened clinical applications. For more than 20 years, early feeding after PEG has been proven safe and widely encouraged [22–23]. However, most evidence supporting early feeding involves pull-technique PEG insertion rather than push-technique placement [2,24]. Consequently, some clinicians remain cautious about complications and delay feeding initiation for 12–24 hours after the procedure [25].

In this randomized controlled trial, we studied patients who underwent push-technique PEG with an intermittent-drip feeding method, the predominant approach at our institution. This technique is favored because a high proportion of cases involve esophageal or head and neck cancers, for which the pull technique is generally unsuitable. Additionally, gastropexy is routinely performed to secure firm attachment of the PEG tube to the abdominal wall.

Maximal GRV did not differ statistically between the early and delayed groups, aligning with prior comparisons of GRV [1,26–27]. Most patients in both groups had a GRV of 0 mL. Eight patients (5 in the early group and 3 in the delayed group) had measurable GRV, but none had sufficient volume to require interrupting subsequent feeding. A limitation of GRV measurement is that aspiration through the PEG site may not capture the entire residual, because contents are withdrawn only when they contact the anterior gastric wall. We also observed no association between GRV and abdominal distension, nausea, or vomiting. One patient in the early group had a maximal GRV of 200 mL, and another in the delayed group had 260 mL; neither experienced gastrointestinal symptoms. These findings suggest that GRV should not be used in isolation to assess feeding intolerance; rather, it should be interpreted together with clinical symptoms, consistent with prior reports [28–29]. Decisions about subsequent feedings should rely primarily on clinical symptoms and physical assessment.

Postprocedural complications were comparable between groups. All events were Clavien–Dindo grade 1 or 2, and no deaths occurred. Among the 4 grade 2 events, 1 occurred in the early feeding group (pneumonia) and 3 in the delayed group (pneumonia, wound bleeding, and urinary tract infection). These results align with prior studies and clinical guidelines supporting the safety of early feeding, including in push-technique PEG. Hospital stay was significantly shorter in the early feeding group, suggesting that early feeding reduces length of stay by allowing patients to reach full caloric intake sooner. Future studies should evaluate the cost-effectiveness of early feeding. The early feeding group received significantly less intravenous morphine than the delayed group, likely because earlier enteral acetaminophen administration reduced reliance on intravenous opioids.

Wound cultures from the PEG site showed no significant differences in bacterial colonization between the early and delayed groups. Coagulase-negative staphylococci, typical Gram-positive skin flora, were most frequently identified. Gram-negative pathogens, including *Klebsiella pneumoniae*, *Pseudomonas aeruginosa*, and *Acinetobacter baumannii*, were also detected in some cases. Although the recovered organisms were similar to prior reports, their prevalence differed from studies reporting the highest proportion of *Candida* spp colonization at the gastrostomy tube tip [30]. This discrepancy likely reflects our sampling of the peristomal wound rather than the tube tip. Pathogen diversity and colony-forming unit counts also did not differ between groups. Although these microbiologic data could inform antibiotic selection for PEG wound infections, typically caused by Gram-positive bacteria, only 1 PEG wound infection occurred and it did not correlate with a positive culture. Prior studies report PEG infection incidence of 21.2%–29.4% [31–32]. Our patients were typically discharged within 2 days, which may have limited detection of infections not yet clinically apparent by postprocedural day 2.

### Limitations

This single-center study from a tertiary care hospital may limit generalizability to institutions with different practices and patient populations. Although the randomized controlled design is a strength, blinding of participants and providers was not feasible and may have introduced observer bias. Measuring GRV by aspiration through the PEG site may not accurately reflect true intragastric contents, as discussed earlier. Discharge within 1–2 days constrained detection of late-onset complications, including infections that might emerge after discharge, particularly among patients with positive wound culture results. The cohort was dominated by esophageal and head and neck malignancies, potentially limiting applicability to broader PEG populations, especially those with neurologic conditions. Finally, although early feeding shortened hospital stay and reduced opioid use, formal cost-effectiveness analyses are needed to substantiate the economic benefits of early initiation.

## Conclusions

Early feeding within 4 hours after push-technique PEG insertion is safe and does not adversely affect postprocedural GRV, complications, or wound-site bacterial colonization. This protocol shortens hospital stay and reduces the need for intravenous morphine.

## Data Availability

Data cannot be shared publicly because of patient confidentiality and ethical restrictions related to this randomized controlled trial.

https://www.thaiclinicaltrials.org/show/TCTR20220105003

## Abbreviations

GRV: Gastric residual volume
PEG: Percutaneous endoscopic gastrostomy
Spp: pecies

## Acknowledgements

The authors thank Miss Wathanaphirom Mangmee and Miss Chorlada Keatrungarun for assistance with data collection, Dr Chulalak Komontri for assistance with statistical analysis, and Mr David Park for his assistance with language editing.

## Funding statement

This trial was funded by the Routine to Research Unit, Faculty of Medicine Siriraj Hospital, Mahidol University.

## Conflict of Interest Statement

The authors have no conflicts of interest to declare.

## Author Contributions

All authors-Conceptualization

TP, WR, CN, JS, AM-Data curation All authors--Formal analysis

TP, WR, CN, AM-Investigation TP, WR, AM-Methodology

TP, WR, JS, AM-Project administration TP, NS, AT, CP, AM, JS-Resources

TP, WR-Software

TP, NS, AT, CP, CN, JS, AM-Supervision All authors--Validation

TP, WR, CN, JS, AM-Visualization All authors-Writing - original draft

All authors-Writing - Review and editing

## Institutional Review Board Statement

The protocol was approved by the Institutional Review Board of the Faculty of Medicine Siriraj Hospital, Mahidol University (Si-930/2021).

## Informed Consent Statement

This study was conducted in accordance with ethical and data protection regulations. Written informed consent was obtained from all subjects involved in the study.

## Data Availability Statement

Data are available upon reasonable request.

## Notes

### Competing Interest Statement

The authors have declared no competing interest.

### Clinical Trial

TCTR20220105003

### Clinical Protocols

https://www.thaiclinicaltrials.org/show/TCTR20220105003.

### Funding Statement

Yes

### Author Declarations

Institutional Review Board of the Faculty of Medicine Siriraj Hospital, Mahidol University (Si-930/2021)

